# The impact of barcode-assisted medication administration on medication administration errors in non-unit-dose settings: a systematic review

**DOI:** 10.1101/2024.10.06.24314967

**Authors:** Weicong Tan, Gordon Bingham, Erica Tong, Erfan Shakibaei Bonakdeh, Weiqing Wang

## Abstract

**Objective:** This study aims to investigate how barcode-assisted medication administration systems (BCMAs) can affect medication administration errors (MAEs) in non-unit-dose dispensing settings, since unit dose dispensing system can be a confounding variable affecting MAE rates.

**Materials and Methods:** We conducted a systematic review of articles on MEDLINE, EMBASE, EMCARE, CINAHL and Scopus. Studies are meticulously examined to exclude those with unit-dose dispensing setting.

**Results:** We included 4 papers in the review. The categories of MAEs reported among these studies are heterogeneous. 2 studies give weak evidence and 1 study give moderate evidence that BCMA can lower some categories of MAEs. 1 studies gives weak evidence that BCMA increase the wrong administration time error.

**Discussion:** Studies provide weak to moderate evidence that barcode-assisted medication administration can lower certain categories of medication administration errors. However, some reported findings are minimal.

**Conclusion:** More multi-ward multi-hospital studies need to be conducted to provide stronger evidence on BCMA’s impact on MAEs, especially on dosage-related MAEs, in settings without unit dose dispensing systems.

## Introduction

Medication administration is an important part of in-hospital care. However, different categories of errors that harm patients can happen in this process [1]. Medication administration errors have been one of the prevalent risks to patients and a challenge imposed on all medical practitioners across the world. The World Health Organization (WHO) identifies medication administration errors as key contributors to patient safety incidents [2].

Medication administration errors (MAEs) encompass any deviation from the prescribed medication order and institutional healthcare policies [3]. To enhance patient safety and guarantee the fundamental “five rights” (i.e., right time, right dose, right drug, right route, and right patient) [4], along with other specifications in the medication order (such as correct solvent/diluent volume [5], right response [6], and right documentation [7]), numerous interventions have been introduced. One of the outstanding approaches is the barcode-assisted medication administration (barcode-assisted medication administration) workflow. It is a systematic method composed of a series of cross-checking (the nurse’s scanning) of the unique identifier of the patient’s identity (the barcode attached to the patient) and the identifier of the medications (the medication’s identifier on the package) [8]. The medication administration process is stored in electronic medical records for analysis, backtracking, and review [9]. The barcode-assisted medication administration workflow aims to minimise the possibility of errors in the medication administration process. This cross-checking aims to ensure the correct medication is administered to the correct patient in the correct route with the correct dose. Moreover, barcode-assisted medication administration can increase the work efficiency of medication administration, thus ensuring the timeliness of the administration [10].

Unit dose dispensing systems were developed in the 1960s to minimise medication waste and medication administration errors [11]. While unit dose dispensing has been standard practice in the United States and is being adopted in other regions (e.g., Germany [12] and Finland [13]), unit dose dispensing is not yet widely adopted as a standard practice in some other regions or jurisdictions (e.g., the United Kingdom [14] and Australia) as of now. Studies (e.g., Taxis et al. (1999) [12] and Means et al. (1975) [15]) showed that unit dose dispensing can lower the probability of medication administration errors. However, previous systematic reviews on barcode-assisted medication administration’s effect on medication administration safety, e.g., Hutton et al. (2021) [16] did not separate non-unit-dose settings from unit-dose settings.

Therefore, it is necessary to eliminate as many factors as possible to investigate the effect of barcode-assisted medication administration on medication administration errors. To our best knowledge, investigate how barcode-assisted medication administration affects patient safety—more particularly, medication administration error rate—in non-unit dose dispensing settings. By attempting to eliminate confounding interventions (e.g., unit dose dispensing system), we can have a better understanding of how barcode-assisted medication administration can facilitate safer medication administration, thus improving care quality and guaranteeing patient safety. Additionally, Hutton et al. (2020) did not conduct a quality assessment for its included studies [16], which is an important part of a systematic review.

## Method

We adopted the Preferred Reporting Items for Systematic Reviews and Meta-Analyses (PRISMA) [17] protocol to select studies for data extraction. The PRISMA checklist of this systematic review can be found in Supplementary Materials.

### Search Strategy and Entry De-duplication

W.T. initiated a thorough search across five electronic databases — MEDLINE, EMBASE, EMCARE, CINAHL, and Scopus—to identify pertinent studies. Our search focused on articles published in academic journals or conferences. The most recent search was conducted on 01/02/2024. The full search strategy for different databases can be found in Supplementary Materials.

All retrieved entries, including duplicates, were uploaded to Covidence [18], where authors conducted the selection of studies. Covidence conducts an automatic de-duplication of studies, after which a subsequent human-in-the-loop approach is conducted by W.T. to ensure the thorough elimination of any remaining duplicates. Titles, abstracts, publication times and authors’ list are cross-checked to identify duplicate entries.

### Eligibility of the Studies

W.T. and E.S.B. independently undertook the initial screening of entries based on their titles and abstracts to assess the relevance of the studies. Subsequently, the same authors conducted a detailed review of the full-text content of the included entries. Where conflicts occur, W.T. and E.S.B. discuss and resolve the conflicts in the title and abstract screening and full-text screening.

To ensure the inclusion of only studies in non-unit-dose settings, the entire authors’ team meticulously examined articles using the following steps:

- **Unit-dose workflow identification**. We carefully examine how the study describes the medication dispensing and administration process. If it is a unit-dose dispensing setting, we exclude this study.
- **Previous studies regarding the dispensing system of the setting**. If it has not been described in the paper whether the study was in a unit-dose setting, the authors search for previous studies regarding its medication dispensing system.
- **Other exclusion protocols**. If no previous studies about the venue dispensing system, the authors discuss the dispensing setting of the study. Factors including the locations of the study are considered while making the final decision.

The inclusion and exclusion criteria for this systematic review are listed as follows:

#### Inclusion Criteria

1. Dispensing setting: non-unit-dose dispensing systems (e.g., Australian, New Zealand and British hospitals)
2. Venue: hospital in-patient wards (N.B. aged care wards in the hospitals are included)
3. Study design: initially we only included studies with a controlled cohort, but limited studies meet this criterion; we then included studies with controlled cohorts, before-and-after studies and quasi-experimental studies
4. Interventions: bedside barcode scanning for patients and medications, implemented by nurses
5. Outcomes: quantitative outcome measuring of the change in medication administration errors

#### Exclusion Criteria

1. Dispensing setting: unit-dose medications dispensing settings (e.g., Canadian and USA hospitals and other hospitals adopting unit-dose dispensing)
2. Venue: medical residential settings that are not hospital wards (e.g., aged care facilities and operation theatres)
3. Non-English papers. We would like to emphasise that this exclusion criteria does not apply in the searching stage of the study.
4. Citations without full text. This exclusion criteria only applies in the full-text review process. We do not check whether a citation has full text available in the title and abstract screening to avoid unnecessary elimination of papers.

### Data Extraction and Quality Assessment

Detailed information is extracted from eligible studies. Key characteristics of included studies are summarised in Table 1.

**Table 1.** Summarisation of key characteristics of included studies.

|  | Franklin et al. (2007) [20] | Franklin et al. (2008) [21] | Barakat et al. (2020) [10] | Westbrook et al. (2020) [5] |
| --- | --- | --- | --- | --- |
| Study design | Observational, before-and-after study | Observational, before-and-after study | Observational, cohort study | Controlled cohort study |
| Quality Assessment | Low, single-site, one ward. | Low, single-site, one ward. | Low, single-site, two wards. | Moderate, multi-site multi-ward controlled cohort study. |
| Setting | A 28-bed general surgery ward in a London teaching hospital, UK. | A 28-bed general surgery ward in a London teaching hospital, UK. | Two surgical wards with 24 beds each, divided into four bays and four side rooms, at a major acute hospital in London, UK. | Two adult teaching hospitals in Sydney, Australia. Four wards in Hospital A and two wards in Hospital B are intervention wards. Two wards in Hospital A and one ward in Hospital B are the control ward. |
| Observers | Pharmacists. | Four pharmacist researchers. | A pharmacy student observed the nurse concerned. | Nurses independent from the study hospitals and underwent extensive training using scenarios and field testing. |
| Intervention | The intervention, launched in June 2003, included a closed-loop system with electronic prescribing, automated dispensing, barcode patient identification, and EMARs (ServeRx V.1:13, MDG Medical, Israel), excluding only intravenous infusions and oral anticoagulants, which remained on paper charts. | A closed-loop system featuring electronic prescribing, automated dispensing, barcode patient identification before drug administration, and eMARs (ServeRx version 1.13, MDG Medical, Tel Aviv, Israel). | The ePMA and barcode-assisted medication administration systems were components of Cerner Millennium. | Both hospitals were implementing a commercial EMS with electronic prescribing and medication administration. Hospital A's intervention wards used Cerner EMS, and Hospital B's intervention ward used iSoft Medchart EMS. |
| No. of Administration | 56 drug rounds. The number of doses was not mentioned. | Pre-intervention: 56 drug rounds and 2336 doses were observed; post-intervention: 55 rounds and 1678 doses were observed | At the non-BCMA ward, 185 administrations were observed. At the BCMA ward, 363 administrations were observed. | 7451 administrations were observed (4176 pre-implementation and 3275 post-implementation). |
| Categories of Errors | Prescribing errors and administration errors. Administration errors include any dose of medication that deviated from the patient's current medication orders (timing and documentation errors were excluded). | Any dose of medication administered deviated from medication orders or omitted doses. | Medications administered at the wrong time. | Wrong intravenous rate, wrong timing, wrong solvent/diluent volume, wrong dose, wrong solvent (for injectables), wrong formulation, wrong strength, IV with incompatible drug, wrong drug, extra dose, wrong route and unordered drug. |

We follow the GRADE Handbook [19] to assess the quality of the eligible studies. The GRADE Handbook is a system to assess the quality of bodies of evidence in the studies in systematic reviews. The quality of a body of evidence is classified into four grades - high, moderate, low and very low. Without strong limitations and special strengths, bodies of evidence provided by randomised trials are given “high” ratings, and bodies of evidence provided by observational studies are given “low” ratings. Ratings can be modified based on various factors, such as limitations, population and imprecision of the study.

### Synthesising the Findings of Included Studies

Any violation in medication administration that violates the fundamental five rights (i.e., right time, right dose, right drug, right route, and right patient) constitutes a medication administration error. Different healthcare institutions have different criteria for a correct medication administration. Therefore, different studies report different categories of medication administration errors. Due to the heterogeneity of outcome metrics, we consider meta-analyses unsuitable for this study. Instead, we use the narrative synthesis approach to summarise the findings of the included studies.

## Results

Figure 1 illustrates the PRIMA flow chart detailing the systematic review process. Initially, 995 studies were retrieved from the five aforementioned databases. Automated processes in Covidence and a human-in-the-loop approach, involving cross-checking authors’ lists, titles, and abstracts, led to the removal of 93 duplicates. Subsequently, the remaining 902 papers underwent screening based on their titles and abstracts. Out of these studies, 788 were excluded as their titles and abstracts met the predetermined exclusion criteria. The remaining 114 papers underwent a thorough review based on their full texts. 2 studies were excluded due to the unavailability of the full text in English, while 27 entries were excluded as their full texts were inaccessible (only the title and/or abstract was available). Additionally, 14 studies were excluded due to their inappropriate setting (e.g., unit-dose dispensing and aged-care facilities), 56 studies were excluded due to their wrong study design, and 1 study were excluded due to the wrong intervention.

**Fig. 1:**
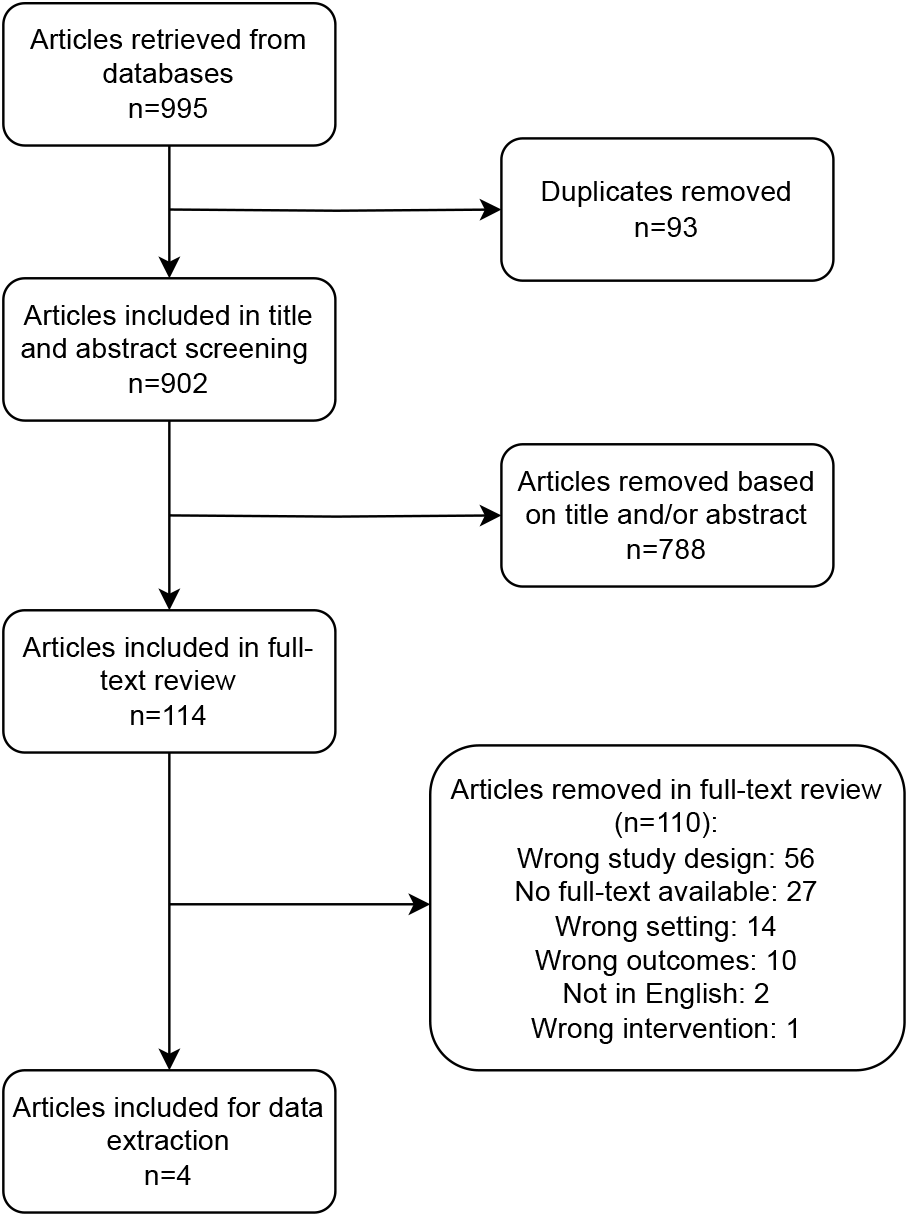
The PRIMA flow chart illustrating the article selection process

Table 1 presents the characteristics of the four included papers, listed in chronological order.

### Study Designs and Settings

Among the four studies included for data extraction, three (Franklin et al. (2007), Franklin et al. (2008) and Barakat et al. (2020)) were located in London, the United Kingdom. One (Westbrook et al. (2020) [5]) was located in Sydney, New South Wales, Australia.

The studies included for this analysis, although not randomised controlled trials, were structured as before-and-after cohort studies. Notably, the study by Westbrook et al. (2020) was a controlled cohort study involving two hospitals. Hospital A involved four general medical/surgical wards with two acute aged care as intervention wards, one renal/ vascular/ dermatology and one acute respiratory ward as control wards. Hospital B involved an acute orthopaedic ward as the intervention ward and an acute neurological ward as the control ward. Barakat et al. (2020) conducted a before-and-after study with two distinct cohorts. Specifically, the pre-intervention group comprised a ward without a barcode-assisted medication administration system, yet sharing a similar physical layout with the post-intervention group, albeit with other uncontrolled variables. Similarly, the studies by Franklin et al. (2007) and Franklin et al. (2008) were both before-and-after investigations conducted within a single cohort, where the interventions were applied to either a singular ward or the same group of wards.

### Interventions of Selected Studies

All studies but one (Barakat et al. (2020)) implemented barcode-assisted medication administration as one of the components in the entire closed-loop medication management system. In the setting of Barakat et al. (2020), the roll-out of barcode-assisted medication administration is incremental. The before-group was a ward with electronic prescribing and administration systems but without a barcode-assisted medication administration system, whereas the after-group was deployed with a barcode-assisted medication administration system. They aim to investigate how barcode-assisted medication administration, as a single component of the entire closed-loop medication management system, can impact the nursing workflow thus the timeliness of medication administration.

The intervention in Franklin et al. (2007) and Franklin et al. (2008) is similar. They are both an entire closed-loop medication management system with an electronic prescribing system, ward-based automated dispensing cabinets, barcode verification of patients’ identity and an electronic medical record system that documents all the medical history. The intervention cohort had been implementing the paper-based medication ordering workflow where doctors prescribed with formatted paper and medications were stored in drug trolleys and stock cupboards.

Westbrook et al. (2020) conducted a controlled cohort study across two hospitals, both in the midst of implementing electronic closed-loop medication management systems. Hospital A adopted the Cerner electronic medication system, whereas Hospital B implemented the iSoft Medchart system. In both hospitals, control groups continued using paper-based medication charts as the standard administration workflow. The intervention involved the deployment of an electronic medication system with electronic provider order entry (ePOE) functionality, enabling clinicians such as doctors, pharmacists, and nurses to enter and access digital notes and documents within an electronic portal.

### Categories of Medication Error Reported

All the included studies investigate at least one type of violation of the five rights. All studies but Barakat et al. (2021) solely aimed to investigate how barcode-assisted medication administration systems affect medication errors. Barakat et al. (2021) aimed to examine the impact of barcode-assisted medication administration on nurses’ activity and workflow [10]. Since barcode-assisted medication administration systems change the nursing workflow, and, therefore, the duration of drug rounds, investigating the timeliness of medication administrations is part of the result of this study.

Westbrook et al. (2020) investigated the impact of barcode-assisted medication administration on medication administration errors. This included both clinical errors (e.g., the violation of the “five rights”) and procedural errors (e.g., the nurse was not following standard medication administration protocol with barcode-assisted systems). In this systematic review, we only analyse the results of clinical errors since these are the metrics directly related to patients’ safety.

Franklin et al. (2007) and Franklin et al. (2008) have the results of both administration errors and prescribing errors, since, as mentioned in 3.2, the ePOE is one of the components of their interventions. Franklin et al. (2007) investigated the violation of the “five rights”, administering extra doses, administering an expired drug, omission of doses (due to unavailability of the drug), omission of doses due to other reasons, wrong diluent and the fast administration of IV bolus. The number of each of these errors is listed before and after the intervention.

However, Franklin et al. (2008) described medication administration errors as any deviation from the original medication order but did not list the specific categories of medication administration errors except the timeliness of the ordered administration. On the contrary, it analysed administration errors for different stocking statuses of drugs. MAEs with ward-stock drugs and non-ward-stock drugs and the omission of doses in these stocking statuses excluding those due to unavailability are investigated. As per the timeliness of the administration, this study listed the before and after data of the difference between the ordered and actual administration time of medication.

### Findings of the Studies

The result of Franklin et al. (2007) showed that the barcode-assisted medication administration technique reduced the number of many categories of administration errors. Wrong drug errors dropped from 2 to 0; wrong dose errors dropped from 29 to 5; wrong patient errors dropped from 5 to 0; wrong route error increased from 2 to 6; wrong time errors increased from 0 to 1; extra dose errors dropped from 2 to 0; administering expired drugs error dropped from 1 to 0; dose omission due to drug unavailability dropped from 26 to 25; dose omission due to all other reasons dropped from 42 to 11; wrong diluent error dropped from 1 to 0; fast administration of IV bolus error dropped from 31 to 5. The total number of administration errors dropped from 141 (which was 8.6% of all opportunities of error) to 53 (which was 4.4% of all opportunities of errors).

Franklin et al. (2008) found out that barcode-assisted medication administration systems decrease the number of MAEs of drugs of different stocking statuses and the timeliness of medication administration increased. MAEs of ward-stock drugs dropped from 77 to 22; MAE for non-ward-stock drugs dropped from 65 to 31; MAE for ward-stock drugs except those due to drug unavailability dropped from 74 to 21; MAE for non-ward-stock drugs except those due to drug unavailability dropped from 41 to 8. The medication administered within one hour from the ordered time dropped from 1719 to 1475; the medication administered within two hour but more than one hour from the ordered time dropped from 1719 to 1475; the medication administered more than two hours from the ordered time dropped from 47 to 0.

Westbrook et al. (2020) found that barcode-assisted medication administration systems (BCMA) reduced certain categories of medication administration errors. In the intervention wards, where BCMA systems were deployed, the rates of several categories of errors were lower compared to control wards. The change in error rates were calculated using the formula [5]:

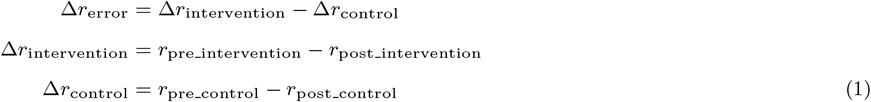

Based on this calculation, compared to control wards, the intervention wards recorded a reduction in wrong intravenous rate errors by 5.7 (per 100 administrations, the same applies below), wrong timing errors by 3.4, wrong dosage errors by 0.4, wrong formulation errors by 0.2, wrong strength errors by 1.1, and extra dose errors by 0.2. However, there was an increase in wrong solvent volume errors for injectables by 4.3, and intravenous administration with incompatible drugs by 0.03, wrong drug errors by 0.3, and wrong route errors by 0.3.

However, Barakat et al. (2020) found that administrations tend to be more timely in the non-BCMA cohort. At the non-BCMA ward, 165 out of 185 (89%) ordered medication were administered; whereas in the BCMA ward, 294 out of 363 (81%) ordered doses were administered. The time difference between ordered time and actual administration time is 60.3 minutes in non-BCMA wards and 67.5 minutes in the BCMA ward. The author argued that the discrepancy is due to the difference in the starting time of drug rounds, and this discrepancy can be possibly offset if the starting times in both cohorts are aligned [10].

### Quality Assessment

None of the eligible studies are randomised trials. Therefore, the initial GRADE [19] ratings of all included studies are “low”. Westbrook et al. (2022) is a controlled study with controlled wards in two different hospitals. Therefore, its quality is modified to “moderate”. However, the authors mentioned that the intervention wards are not randomly selected due to operation reasons. Franklin et al. (2007), Franklin et al. (2008) and Barakat et al. (2020) all received “low” quality ratings due to their lack of control groups and small population.

## Discussion

### Summary of Findings and Implications

The findings of most included studies suggested that, in a non-unit dose dispensing setting, barcode-assisted medication administration systems can reduce medication administration error rates to some degree. One study (Westbrook et al. (2020) [5]) gave moderate evidence that barcode-assisted medication administration systems can lower some reported categories of medication administration errors, and two studies gave weak evidence (Franklin et al. (2007) [20] and Franklin et al. (2008) [21]).

One of the objectives of this study is to investigate the impact of barcode-assisted administration systems on dosage-related medication administration rates in non-unit-dose settings. Only two studies (i.e., Franklin et al. (2007) [20] and Westbrook et al. (2020) [5]) reported the change in wrong dosage errors. While Franklin et al. (2007) reported a relatively larger decrease in dosage-related error, Westbrook et al. (2020) reported small decreases in some dosage-related errors (wrong dose, wrong strength and wrong formulation). With the included studies, we only have weak evidence that, in non-unit-dose settings, barcode-assisted administration systems can lower the rate of dosage related medication administration errors.

For wrong timing administration errors, Franklin et al. (2008) and Westbrook et al. (2020) reported that the introduction BCMA reduced wrong timing administration errors, whereas Barakat et al. (2020) and Franklin et al. (2007) reported the opposite result. The number of wrong timing errors in Franklin et al. (2007) rose from 0 to 1, which is minimal. However, the increase in the number of timing errors was larger in Barakat et al. (2020), which may be due to two reasons. Firstly, Barakat et al. (2020) had a more broad definition of “administration timeliness”, which included patients’ refusals, the unavailability of the medication, and the need for reviews in this administration. Secondly, in the setting of Barakat et al. (2020), the time of the drug rounds are different in BCMA and non-BCMA wards.

All included studies are not randomised controlled studies. It is worth noticing that it is extremely difficult to conduct randomised studies since it is difficult to do so in operating healthcare institutions. However, two studies made efforts to introduce randomness. Barakat et al. (2020) randomly selected the nursing bay to be observed every day [10], and Westbrook et al. (2020) randomly selected consented nurses to observe every day [5]. One way to give stronger evidence is to involve more wards and sites in a study. However, Westbrook et al. (2020) [5] did not report the separate results in the two involved hospitals. This imposes a risk of bias since the difference in wards or hospitals may be a confounding variable affecting the rate of medication administration error, especially when the two hospitals in Westbrook et al. (2020) [5] adopted different electronic systems.

It is worth noticing that for all included studies, the data collection process for post-intervention started at a certain period of time after the deployment of the system. This ensures the results can reduce the recording of errors caused by the errors made by the nurses while adapting to the new system.

Included studies were conducted in two hospitals in London, the United Kingdom and two hospitals in Sydney, Australia. These are major cities in countries with high socio-economic status. It indicates that we need more high-quality research investigating the impact of barcode-administration medication administration systems on medication administration errors in regional hospitals and hospitals in other countries, especially countries with relatively lower socio-economic status, where the barcode-assisted medication administration systems may be gradually adopted.

### Strengths and Limitations

Several systematic reviews have been conducted to summarise the impact of barcode technology on medication errors, including Hutton et al. (2021) [16]. However, Hutton et al. (2021) [16] summarised results in both unit-dose and non-unit-dose settings and lacked quality/risk of bias assessment. In our study, we conduct quality assessment on included studies, which is an integral component of a systematic review. Moreover, we synthesise only results in non-unit-dose settings, which excludes an important confounding variable that causes medication administration errors, especially dosage errors.

However, our study has several limitations. Due to the heterogeneity of medication administration errors, it is impractical to conduct meta-analyses. This systematic review is based on a relatively small number of studies, especially controlled studies, due to our elimination of studies in unit dose settings.

## Conclusion

In this systematic review, we summarise four studies that investigated the impact of the barcode-assisted medication administration technique in a non-unit-dose dispensing setting on medication administration errors. The summarised findings provide weak evidence that barcode-assisted medication administration can reduce some categories of medication administration errors. According to the quality assessment of this study, due to the lack of high-quality research with randomised trials, it is difficult to draw a conclusive statement that the barcode-assisted medication administration can significantly improve patient safety in a non-unit-dose dispensing setting. One low-quality study also suggested that barcode-assisted medication administration cannot lower the rate of wrong administration time error [10]. Due to the operational difficulty of conducting randomised controlled studies, future evaluations of its impact need to be conducted by controlled studies with more wards and hospitals, and the results of these wards and hospitals should be reported separately, which can provide stronger evidence of the impact of barcode-assisted medication administration on medication administration errors.

## Supporting information

PRIMA checklist

Search strategy for MEDLINE

Search strategy for EMBASE and EMCARE

Search strategy for CINAHL

Search strategy for Scopus

LaTeX souce file

## Data Availability

Not applicable.

## Author contributions statement

W.T. conducted the searching and screening of studies, analysed the data, and wrote the manuscript. G.B. and E.T. designed the inclusion and exclusion criteria, participated in the screening of the studies, and reviewed the manuscript. E.S.B. screened the studies as the second reviewer and reviewed the manuscript. W.W. reviewed the manuscript.

## Acknowledgements

W.T. is supported by the Graduate Research Industry Partnership (GRIP) PhD scholarship, jointly funded by Alfred Health and Monash University. While participating in this research project, E.S.B was a PhD candidate supported by the GRIP PhD scholarship.

