## Supplementary figures and images for "The impact of barcode-assisted medication administration on medication administration errors in non-unit-dose settings: a systematic review"

### PRIMA.pdf

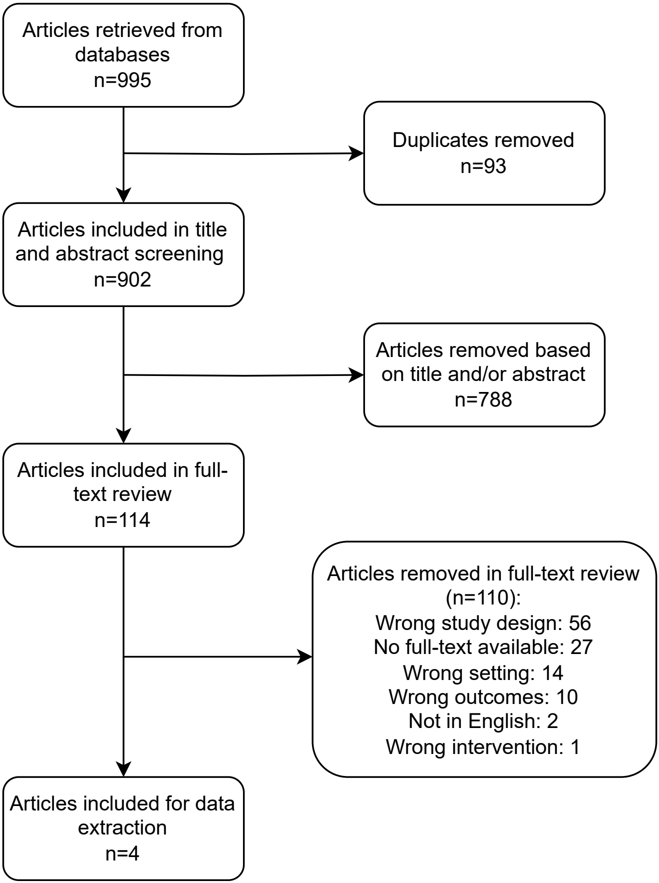

### Search strategy for CINAHL

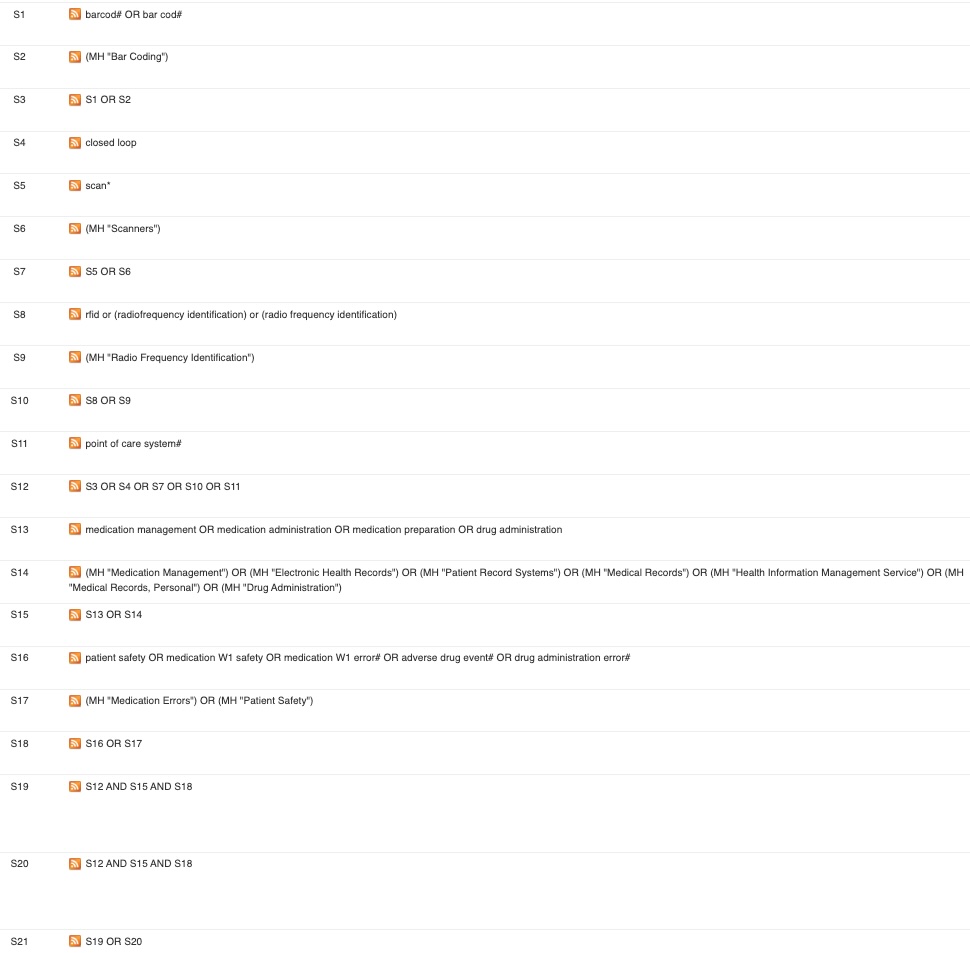

### Search strategy for EMBASE and EMCARE

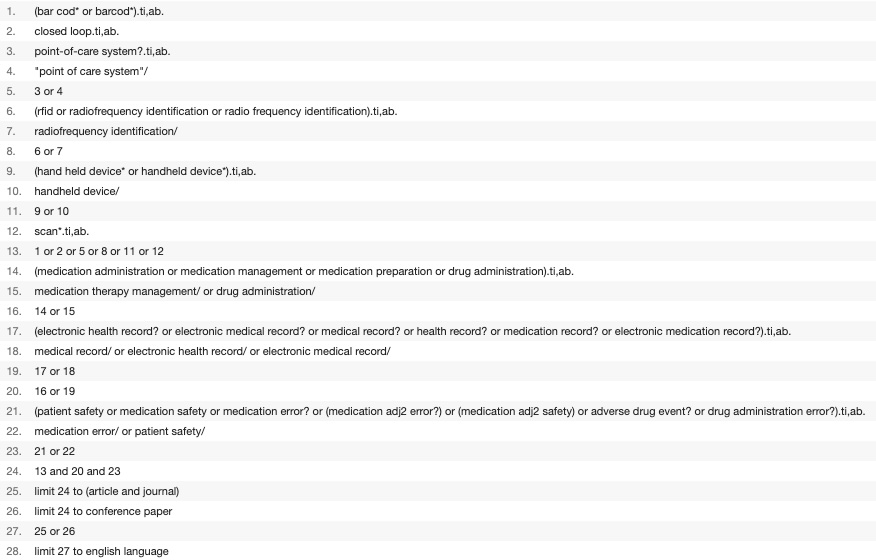

### Search strategy for MEDLINE

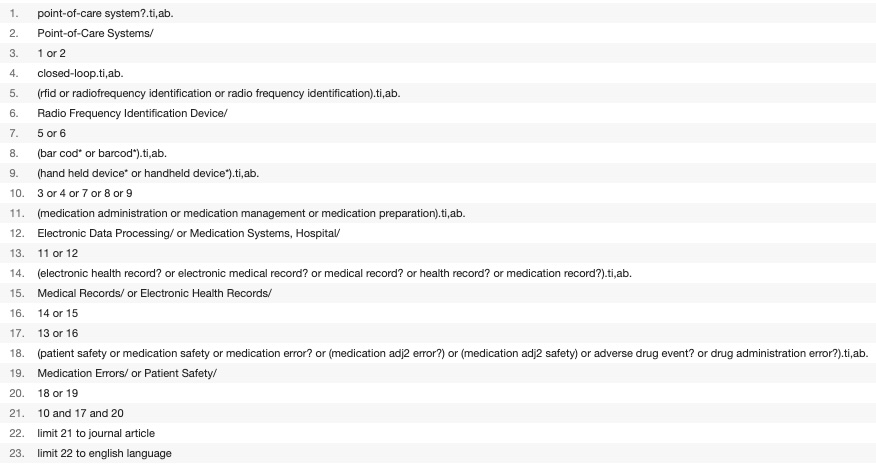

### Search strategy for Scopus

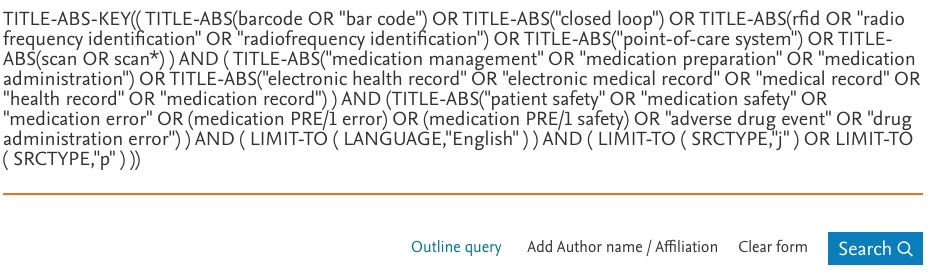
