## Supplementary material for "The impact of barcode-assisted medication administration on medication administration errors in non-unit-dose settings: a systematic review": LaTeX souce file: reference.bib

@article{murray2001unit,
  title={Unit-dose drug distribution systems},
  author={Murray, Michael D and Shojania, Kaveh G},
  journal={Making health care safer: a critical analysis of patient safety practices},
  volume={101},
  year={2001},
  publisher={Citeseer}
}

@article{poon2010effect,
  title={Effect of bar-code technology on the safety of medication administration},
  author={Poon, Eric G and Keohane, Carol A and Yoon, Catherine S and Ditmore, Matthew and Bane, Anne and Levtzion-Korach, Osnat and Moniz, Thomas and Rothschild, Jeffrey M and Kachalia, Allen B and Hayes, Judy and others},
  journal={New England Journal of Medicine},
  volume={362},
  number={18},
  pages={1698--1707},
  year={2010},
  publisher={Mass Medical Soc}
}

@article{young2010bar,
  title={Bar code technology and medication administration error},
  author={Young, Judith and Slebodnik, Maribeth and Sands, Laura},
  journal={Journal of patient safety},
  volume={6},
  number={2},
  pages={115--120},
  year={2010},
  publisher={JSTOR}
}

@article{taxis1999hospital,
  title={Hospital drug distribution systems in the UK and Germany-a study of medication errors},
  author={Taxis, Katja and Dean, Bryony and Barber, Nick},
  journal={Pharmacy World and Science},
  volume={21},
  pages={25--31},
  year={1999},
  publisher={Springer}
}

@article{bint2023developing,
  title={Developing a unit dose approach to managing oral opioid liquids in an Australian hospital},
  author={Bint, Lewis and Rawlins, Matthew and Lawrence, Ashleigh and Tam, Ken and Jenkins, Barry},
  journal={Journal of Pharmacy Practice and Research},
  year={2023},
  publisher={Wiley Online Library}
}

@article{mohanasundaram2021reduction,
  title={Reduction of oral liquid controlled drugs discrepancy in day-to-day practice},
  author={Mohanasundaram, Shalini and Rutter, Paul and Fox, Andrew and Wood, Kelly and Roldo, Marta},
  journal={International Journal of Pharmacy Practice},
  volume={29},
  number={4},
  pages={356--361},
  year={2021},
  publisher={Oxford University Press UK}
}

@article{sinnemaki2014starting,
  title={Starting an automated dose dispensing service provided by community pharmacies in Finland},
  author={Sinnem{\"a}ki, Juha and Saastamoinen, Leena K and Hannula, Sara and Peura, Sirpa and Airaksinen, Marja},
  journal={International journal of clinical pharmacy},
  volume={36},
  pages={345--351},
  year={2014},
  publisher={Springer}
}


--pharmaceutical
@article{catania2011cloperastine,
  title={Pharmacological and clinical overview of cloperastine in treatment of cough},
  author={Catania, Maria Antonietta and Cuzzocrea, Salvatore},
  journal={Therapeutics and clinical risk management},
  volume={7},
  pages={83},
  year={2011},
  publisher={Dove Press}
}

@article{nurse_report,
author = {KIM, KEUM SOON and KWON, SO-HI and KIM, JIN-A and CHO, SUNHEE},
title = {Nurses’ perceptions of medication errors and their contributing factors in South Korea},
journal = {Journal of Nursing Management},
volume = {19},
number = {3},
pages = {346-353},
keywords = {medication errors, nurses’ perception, patient safety, reporting medication errors},
doi = {https://doi.org/10.1111/j.1365-2834.2011.01249.x},
year = {2011}
}

@article{electric_presc_effect,
author = {Shawahna, Ramzi and Ur-Rahman, Nisar and Ahmad, Mahmood and Debray, Marcel and Yliperttula, Marjo and Declèves, Xavier},
year = {2011},
month = {05},
pages = {3233-45},
title = {Electronic prescribing reduces prescribing error in public hospitals},
volume = {20},
journal = {Journal of clinical nursing},
doi = {10.1111/j.1365-2702.2011.03714.x}
}

@article{ozturk2020effect,
  title={Effect of Closed Loop Medication Administration on Drug Returns in Inpatient Facilities},
  author={Ozturk, Eceberil and Kose, Ilker and Elmas, Beytiye Ozge},
  journal={Medical Research Archives},
  volume={8},
  number={12},
  year={2020}
}

@article{hutton2021effects,
  title={The effects of bar-coding technology on medication errors: a systematic literature review},
  author={Hutton, Kevin and Ding, Qian and Wellman, Gregory},
  journal={Journal of Patient Safety},
  volume={17},
  number={3},
  pages={e192--e206},
  year={2021},
  publisher={LWW}
}

@article{Westbrook2020,
   author = {Westbrook, J. I. and Sunderland, N. S. and Woods, A. and Raban, M. Z. and Gates, P. and Li, L.},
   title = {Changes in medication administration error rates associated with the introduction of electronic medication systems in hospitals: a multisite controlled before and after study},
   journal = {BMJ Health Care Inform},
   volume = {27},
   number = {3},
   DOI = {10.1136/bmjhci-2020-100170},
   year = {2020},
   type = {Journal Article}
}

@article{Barakat2020,
   author = {Barakat, S. and Franklin, B. D.},
   title = {An Evaluation of the Impact of Barcode Patient and Medication Scanning on Nursing Workflow at a UK Teaching Hospital},
   journal = {Pharmacy (Basel)},
   volume = {8},
   number = {3},
   DOI = {10.3390/pharmacy8030148},
   year = {2020},
   type = {Journal Article}
}

@article{Franklin2007,
   author = {Franklin, B. D. and O'Grady, K. and Donyai, P. and Jacklin, A. and Barber, N.},
   title = {The impact of a closed-loop electronic prescribing and administration system on prescribing errors, administration errors and staff time: a before-and-after study},
   journal = {Qual Saf Health Care},
   volume = {16},
   number = {4},
   pages = {279-84},
   DOI = {10.1136/qshc.2006.019497},
   year = {2007},
   type = {Journal Article}
}

@article{Franklin2008,
   author = {Franklin, Bryony Dean and Jacklin, Ann and Barber, Nick},
   title = {The impact of an electronic prescribing and administration system on the safety and quality of medication administration},
   journal = {International Journal of Pharmacy Practice},
   volume = {16},
   number = {6},
   pages = {375-379},
   DOI = {10.1211/ijpp.16.6.0006},
   year = {2008},
   type = {Journal Article}
}

@article{RN1698,
   author = {Noguchi, Chihiro and Sakuma, Mio and Ohta, Yoshinori and Bates, David W. and Morimoto, Takeshi},
   title = {Prevention of Medication Errors in Hospitalized Patients: The Japan Adverse Drug Events Study},
   journal = {Drug Safety},
   volume = {39},
   number = {11},
   pages = {1129-1137},
   abstract = {The nature of medication errors (MEs) and the frequency of identified or intercepted MEs are not being scrutinized in daily practice in Japan.},
   ISSN = {1179-1942},
   DOI = {10.1007/s40264-016-0458-1},
   year = {2016},
   type = {Journal Article}
}

@article{tolley2022impact,
  title={The impact of a novel medication scanner on administration errors in the hospital setting: a before and after feasibility study},
  author={Tolley, Clare L and Watson, Neil W and Heed, Andrew and Einbeck, Jochen and Medows, Suzanne and Wood, Linda and Campbell, Layla and Slight, Sarah P},
  journal={BMC Medical Informatics and Decision Making},
  volume={22},
  number={1},
  pages={1--9},
  year={2022},
  publisher={BioMed Central}
}

@article{koyama2020effectiveness,
  title={Effectiveness of double checking to reduce medication administration errors: a systematic review},
  author={Koyama, Alain K and Maddox, Claire-Sophie Sheridan and Li, Ling and Bucknall, Tracey and Westbrook, Johanna I},
  journal={BMJ quality \& safety},
  volume={29},
  number={7},
  pages={595--603},
  year={2020},
  publisher={BMJ Publishing Group Ltd}
}

@article{means1975medication,
  title={Medication errors in a multidose and a computer-based unit dose drug distribution system},
  author={Means, Berry J and Derewicz, Henry J and Lamy, Peter P},
  journal={American Journal of Health-System Pharmacy},
  volume={32},
  number={2},
  pages={186--191},
  year={1975},
  publisher={Oxford University Press}
}

@article{jessurun2022cost,
  title={Cost-effectiveness of central automated unit dose dispensing with barcode-assisted medication administration in a hospital setting},
  author={Jessurun, Janique Gabri{\"e}lle and Hunfeld, Nicole Geertruida Maria and van Dijk, Monique and Polinder, Suzanne and others},
  journal={Research in Social and Administrative Pharmacy},
  volume={18},
  number={11},
  pages={3980--3987},
  year={2022},
  publisher={Elsevier}
}

@article{conroy2007interventions,
  title={Interventions to reduce dosing errors in children: a systematic review of the literature},
  author={Conroy, Sharon and Sweis, Dimah and Planner, Claire and Yeung, Vincent and Collier, Jacqueline and Haines, Linda and Wong, Ian CK},
  journal={Drug Safety},
  volume={30},
  pages={1111--1125},
  year={2007},
  publisher={Springer}
}

@article{kim2018errors,
  title={Errors associated with the rights of medication administration at hospital settings},
  author={Kim, PC and Sen, JJ and Angosta, AD and Frakes, K and Li, C},
  journal={Journal of Hospital and Healthcare Administration},
  year={2018}
}

@article{elliott2010nine,
  title={The nine rights of medication administration: an overview},
  author={Elliott, Malcolm and Liu, Yisi},
  journal={British Journal of Nursing},
  volume={19},
  number={5},
  pages={300--305},
  year={2010},
  publisher={MA Healthcare London}
}

@article{tsegaye2020medication,
  title={Medication administration errors and associated factors among nurses},
  author={Tsegaye, Dejene and Alem, Girma and Tessema, Zenaw and Alebachew, Wubet},
  journal={International journal of general medicine},
  pages={1621--1632},
  year={2020},
  publisher={Taylor \& Francis}
}

@article {Keerse005948,
	author = {Richard N Keers and Steven D Williams and Jonathan Cooke and Darren M Ashcroft},
	title = {Understanding the causes of intravenous medication administration errors in hospitals: a qualitative critical incident study},
	volume = {5},
	number = {3},
	elocation-id = {e005948},
	year = {2015},
	doi = {10.1136/bmjopen-2014-005948},
	publisher = {British Medical Journal Publishing Group},
	issn = {2044-6055},
	eprint = {https://bmjopen.bmj.com/content/5/3/e005948.full.pdf},
	journal = {BMJ Open}
}
	URL = {https://bmjopen.bmj.com/content/5/3/e005948},


@article{jessurun2023prevalence,
  title={Prevalence and determinants of medication administration errors in clinical wards: a two-centre prospective observational study},
  author={Jessurun, Janique Gabri{\"e}lle and Hunfeld, Nicole Geertruida Maria and de Roo, Michelle and van Onzenoort, Hein Antonius Walterus and van Rosmalen, Joost and van Dijk, Monique and van den Bemt, Patricia Maria Lucia Adriana},
  journal={Journal of Clinical Nursing},
  volume={32},
  number={1-2},
  pages={208--220},
  year={2023},
  publisher={Wiley Online Library}
}

@article{who2021global,
  title={Global patient safety action plan 2021--2030: towards eliminating avoidable harm in health care},
  author={World Health Organization and others},
  year={2021},
publisher={World Health Organization}
}


@software{covidence,
  author = {{Veritas Health Innovation}},
  title = {Covidence systematic review software},
  url = {www.covidence.org},
}

@misc{prisma,
  title={Preferred reporting items for systematic reviews and meta-analyses: The PRISMA statement. Annals ofInternal Medicine, 151 (4), 264-269},
  author={Moher, D and Liberati, A and Tetzlaff, J and Altman, DG},
  year={2009}
}

@misc{grade,
  title={The GRADE handbook},
  author={Sch{\"u}nemann, Holger and Bro{\.z}ek, Jan and Guyatt, Gordon and Oxman, Andrew and others},
  year={2013},
  publisher={Cochrane Collaboration London, UK}
}

@article{zheng2021impact,
  title={The impact of introducing automated dispensing cabinets, barcode medication administration, and closed-loop electronic medication management systems on work processes and safety of controlled medications in hospitals: A systematic review},
  author={Zheng, Wu Yi and Lichtner, Valentina and Van Dort, Bethany A and Baysari, Melissa T},
  journal={Research in Social and Administrative Pharmacy},
  volume={17},
  number={5},
  pages={832--841},
  year={2021},
  publisher={Elsevier}
}
